# Characterizing the connection between Parkinson’s disease progression and healthcare utilization

**DOI:** 10.1101/2024.09.15.24313708

**Authors:** Lane Fitzsimmons, Francesca Frau, Sylvie Bozzi, Karen J. Chandross, Brett K. Beaulieu-Jones

**Author notes:** Corresponding author, 5841 S. Maryland Avenue, MC 6092 Chicago, IL 60637-1470.

## Abstract

**Background and Objectives:** Parkinson’s disease (PD) progression can be characterized in terms of healthcare utilization by analyzing clinical events across different stages of disease.

**Methods:** PD progression was measured by the Hoehn & Yahr (H&Y) clinical rating scale and clinical events at each stage were evaluated. Natural language processing and a large language model were used to extract H&Y values from real-world data enabling a larger cohort than manually collected studies, and multi-state hidden Markov models were used for H&Y progression likelihood.

**Results:** Within the one year, most patients in H&Y stages 2-5 remained in the same stage. Stage transitions, when they occurred, were most frequently to the next higher stage. Higher H&Y stages were associated with discharges into long term care and higher rates of additional clinical events.

**Conclusions:** Stratifying key clinical events by H&Y score demonstrates the increases of health care utilization and economic burden with PD severity. Modelling the progression likelihood establishes a progression timeline and emphasizes the unmet need to identify treatment options that stop or slow these transitions.

## Introduction

As Parkinson’s disease (PD) progresses and symptoms become more severe, a patient’s ability to perform daily activities may substantially deteriorate. Patients with advanced PD may not be able to maintain their independence and eventually require specialized support in long-term care facilities. As PD advances, the increasing healthcare utilization required presents a substantial cost. The total economic burden of the disease is an estimated $51.9 billion, with direct medical expenses responsible for $25.4 billion [1].

PD progresses gradually and clinicians use The Hoehn and Yahr Scale (H&Y) to quantify symptom severity [2]. Patients are scored on a scale of 1 to 5, with 1 indicating mild onset of symptoms and 5 indicating severe impairment. The progression through these stages is not linear, and patients may advance at different rates or potentially skip stages entirely [3]. In real-world datasets, structured H&Y score data is not regularly available. Manually analyzing free clinical text and assigning H&Y scores is arduous and difficult to scale. The use of natural language processing and large language models in extracting these values enables the feasible, quantifiable assessment of score transitions in real-world datasets at scale.

Understanding the frequency of progression from one H&Y stage to another would support managing appropriate treatment plans and expectations for individuals with PD. Associating clinical events and health care utilization data with H&Y stages would develop the characterization of a stage in terms of quality of life and economic burden. Using both study level and real-world data, we characterize H&Y scores in terms of health care utilization and evaluate the likelihood of progressing from one H&Y stage to another. This characterization of PD progression and its connection to healthcare utilization can provide an understanding of the potential value of both therapeutic and other treatments aiming to treat PD and/or slow disease progression.

## Results

This study uses two primary sources of data originally described in Beaulieu-Jones et al.[4], electronic medical records (EMRs) from Mass General Brigham (MGB) in addition to a longitudinal cohort study, the Harvard Biomarker Study (HBS).Because most real-world data, including MGB and HBS, are irregularly sampled, it is not straightforward to use these raw observations directly to assess typical progression. To convert from the irregularly sampled H&Y measurements that originate from both passively collected EMRs (MGB) and a registry with irregular follow-up patterns (HBS), we modelled transition likelihood using multi-state hidden Markov models (Figure 1). Using irregularly sampled data as an input, this allowed for an estimate of progression on a fixed interval.

**Figure 1.**
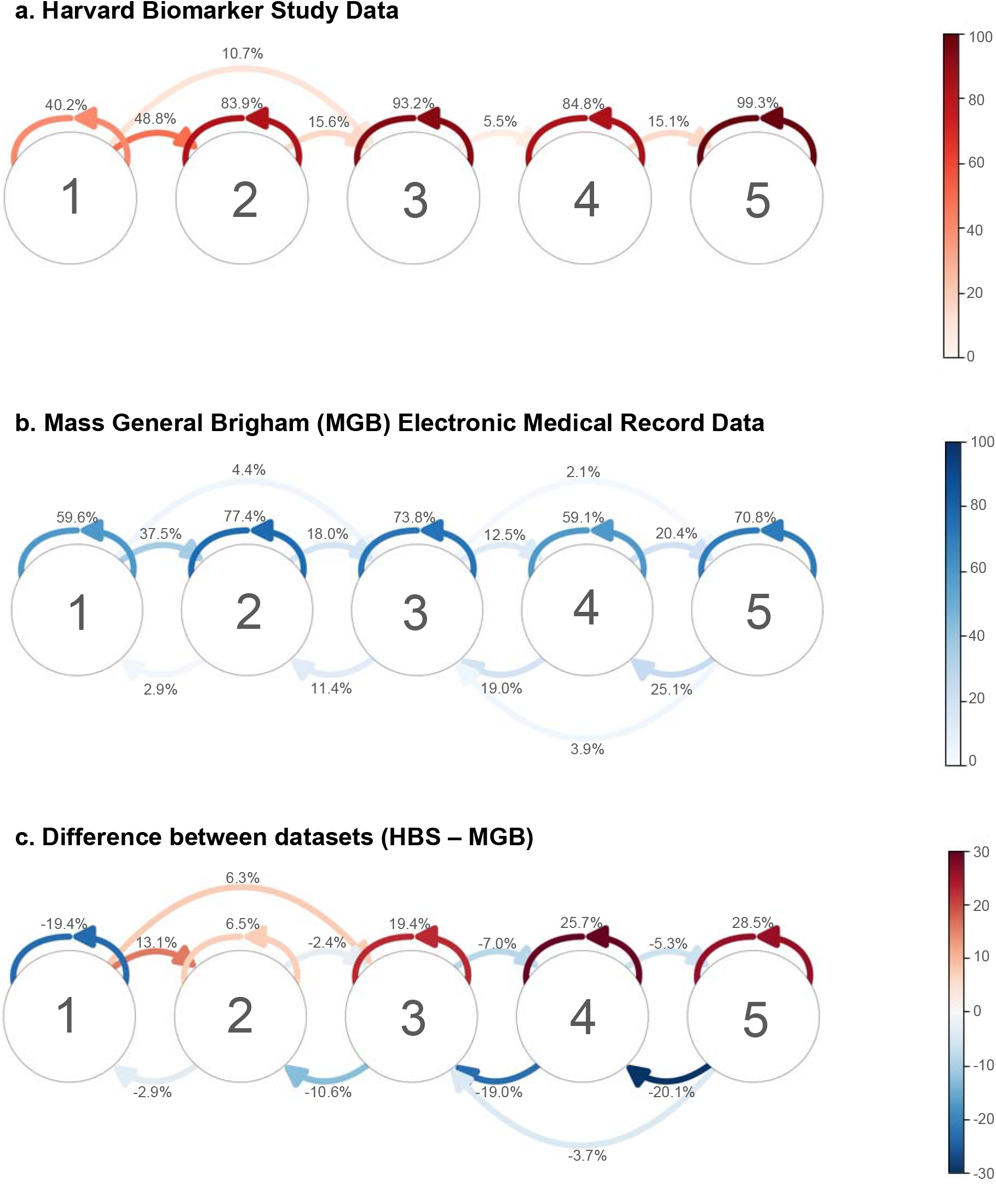
Modelled 12-month H&Y transition likelihood using Multi-state Hidden Markov Models of in Continuous Time. **a.)**, Harvard Biomarker Study Data. **b.)**, Mass General Brigham (MGB) Electronic Medical Record Data. c.), Difference between datasets (HBS – MGB)

A patient’s movement from one H&Y stage to another comes with significant changes to their impairment and quality of life. Modelling the frequency of this transition allowed us to better understand the PD progresses over a year and even suggested the potential benefit that would come with successful intervention. Figure 1 and Supplemental Table 1 show the modeled likelihood that a person with PD transitioned from one state to the same or another state within one year. Only transitions with values greater than 2.0% are displayed. In a one-year snapshot of the significantly longer PD progression process, the majority of patients who began in stages 2-4 remained in the same stage by the end of the period. In both datasets, the highest frequency of patients transitioning from one stage to another was between stage 1 and 2 (48.8% HBS, 37.5% MGB).

Progression was largely unidirectional. In both datasets, patients who were not already in the final stage of disease changed stages with the highest frequency to the adjacent higher stage. For the HBS dataset, less than 1% of patients at each stage transitioned from a higher to a lower H&Y score. For the noisier MGB dataset, at least 2.9% of patients at each stage experienced transitions in this direction.

We analyzed health care utilization and clinical events associated with PD progression according to H&Y to associate quality of life and economic costs with H&Y (Table 1). Discharge to long-term care, and the rates of falls or fractures, were associated with H&Y score. Overall, these event rates could be used to evaluate the potential value of an experimental treatment to slow progression of PD or other symptomatic treatments which may reduce specific adverse events (e.g., a treatment which reduces falls but does not otherwise effect progression).

**Table 1.**
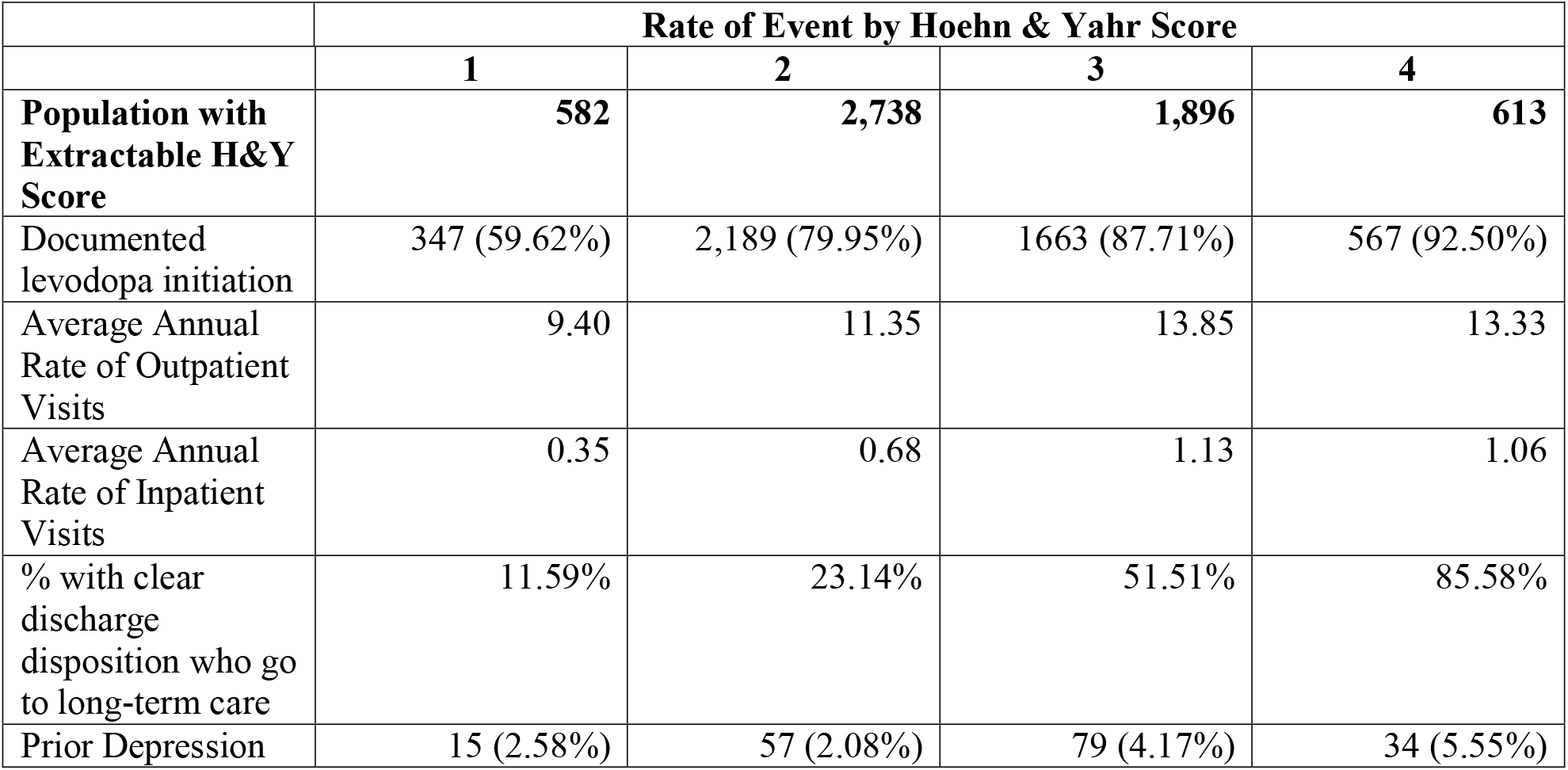
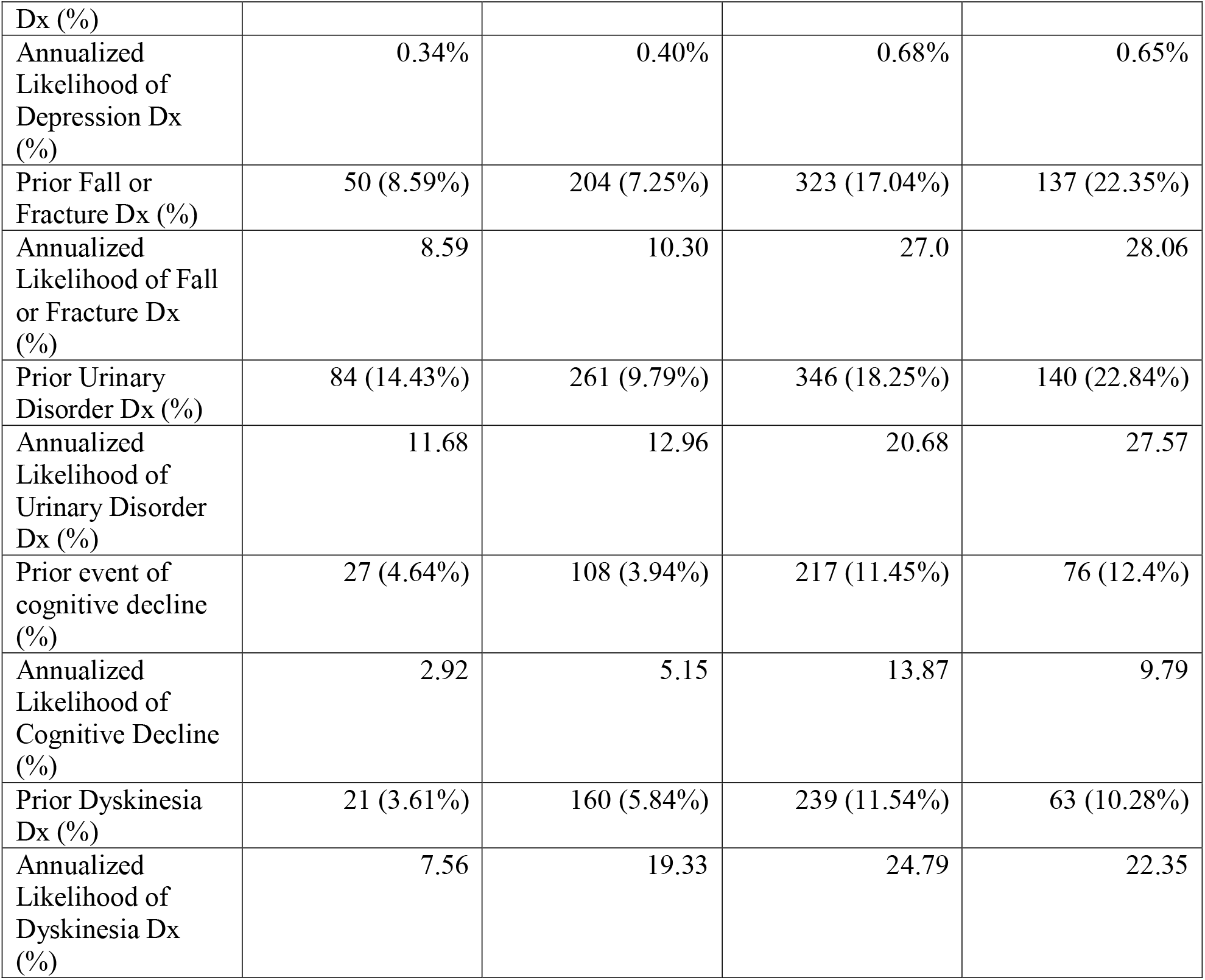
Health Care Utilization and Clinical Events stratified by Hoehn & Yahr Score (MGB).

As H&Y stage increased, patients had higher rates of falls or fractures, urinary disorders, and discharges to long term care. Annual inpatient and outpatient visits, and annualized likelihood of depression, cognitive decline, and dyskinesia trended upward from H&Y 1 to 3, and then downward from H&Y 3 to 4. At H&Y score 4, documented Levodopa initiation reached 92.50%. Data from patients at H&Y stage 5 was not included in Table 1 due to sample size and expert clinical guidance that due to severity of disease at H&Y, the patient condition is obvious enough that tracking and continued recording of H&Y stage was unlikely. H&Y 5 was included in Figure 1 because it allowed for a comparison to the actively collected data for the HBS cohort, but clinical utilization cannot be assessed in the HBS cohort.

## Discussion

This study informs our understanding of PD progression particularly in terms of the connection between adverse events and H&Y scores. It examines both transition likelihood from one H&Y stage to another as well as the characteristics present in patients at each stage. As patients experience gradual progressive decline, their symptom severity is characterized by an increasing H&Y score and decreasing capacity for independence. This study leverages the advances in natural language processing to extract structured information (H&Y scores) from unstructured data (clinical notes) enabling the assessment of H&Y progression and characterization of healthcare utilization in real-world data. This is especially important when considering striking differences between real-world and research populations[4]. Finally, this work leveraged statistical approaches (Hidden Markov Models) to characterize “typical” progression from irregularly sampled data.

Modelling progression over a 12-month period provides of snapshot of PD progression, which in its entirety, may last decades. Because of PD’s slow progression, an extensive time period would be required to assess patients across the span of the H&Y scale. This timeline makes using H&Y scores as an evaluation metric for clinical trials challenging. Patients can remain in one stage for years, so interventions that may result in overall progression abatement may not have results apparent within a reasonable window of study.

The most frequent transition was from stage 1 to stage 2. Consistently, the progression from stage 1 to 2 has previously been found to have the shortest transition period (median 20 months). For all other stages, the majority of patients remaining same stage is consistent with previously documented median transition times for these stages of least 2 years [3]. Fewer of these longer transitions are likely to be captured in our model given the period of just 12-months.

While it is plausible and potentially expected that patients at low H&Y scores (e.g., 1-2) may improve upon the initiation of therapeutics, particularly levodopa, it is not expected that patients at high H&Y scores would show significant improvement. Within the HBS dataset, improvement at high H&Y starting points was rare, but we observed more noise in the MGB data. We attribute this to the fact that these data are collected as part of standard clinical care, and it is not possible to guarantee that a H&Y value recorded in a note means that H&Y was assessed in that specific visit (i.e., prior scores can be copy and pasted to future visits etc.). This highlights an important potential limitation in using data collected for the purposes of standard clinical care for other purposes in analyses of these types despite attempts to filter incorrectly copied scores (e.g., automated detection of incorrect dates).

To assess impacts on quality of life and healthcare utilization, this work also examines clinical factors associated with each H&Y stage. Patients require greater medical care as their H&Y stage increases. At stages 3 and 4, rates of depression, cognitive decline, urinary disorders, and dyskinesia were higher than those at stages 1 and 2. Increases in these clinical events such as depression, cognitive decline, urinary disorders, and dyskinesia do not only negatively impact patients’ quality of life themselves, but also can also exacerbate additional risk of injury. Cognitive impairment, dyskinesia, and antidepressant use have been found to be associated with falls [5]. Consistently, we found that as H&Y stage increased, patients had higher rates of PD-associated adverse events like falls and fractures.

We did also observe that within the EMR data and particularly at higher scores, there is a small percentage of movement to lower scores. We believe this could potentially be due to therapeutic state (e.g., score when “on” or “off” of levodopa or before and after the initiation of therapy), in addition to small amounts of measurement error particularly when as the H&Y score rises, the importance and utility of tracking it degrades (i.e., it is important to track prior to H&Y 3, less so at 3 and above). The combination of clinical events can interfere with patients’ ability to perform everyday tasks and eventually threaten their independence. As H&Y stage increases, patients are more frequently discharged into long term care. With the availability of facility healthcare, hospital visits might become less necessary, and changes in patients’ PD progression may be evaluated less frequently. This is consistent with the slight downward trend of some elements of healthcare utilization after H&Y stage 3.

Using health care utilization as a proxy for economic burden, the substantial cost of PD medical care demonstrates the potential value of limiting its development. A robust understanding of PD progression is critical to managing expectations and created informed treatment plans for patients. Characterizing H&Y scores in terms of progression likelihood and clinical events contributes to this understanding and demonstrates the high value of further PD research.

## Methods

### Data Sources and Study Population

Authors had primary access to all data sources used in this study (BKB). The data sources included in this study are a subset of those used in Beaulieu-Jones et al., where the detailed data descriptions can be found [4]. Cohort characteristics are included in Supplemental Table 2.

### Extraction of Hoehn & Yahr

The process to extract H&Y scores exactly matched the process described in Beaulieu-Jones et al. As previously described “200 notes where the reviewer identified a score, 200 notes where the model identified a score, 200 notes where no score was found by the model, and 200 notes where we manually identified that no score was found. For the 800 notes, extraction accuracy was 99.75%.”

### Study Design and Analysis

Analyses were primarily performed in Python 3.7, with the exception of the use of R for Multi-state Hidden Markov Modeling using the “msm” package [6,7].

#### Statistical Analyses of Extracted Scores (MGB)

As a secondary validation of extracted scores, we compared the median transition time between H&Y stages with registry-reported values from the Zhao et al., reported in Supplementary Table 5 of our prior work, Beaulieu-Jones et al. [3,4]. We expect differences between the scores extracted from MGB and the scores presented in Zhao et al. for two primary reasons: 1.) population differences, MGB comprises a predominately Caucasian population while Zhao et al. report from the movement disorder database at the National Neuroscience Institute in Singapore. 2.) MGB is a passively collected EMR, while Zhao et al. derived data from a registry with regularly scheduled appointments. We expect a passive Electronic Medica Record would report transitions at a slower rate in general. The similarities between MGB and Zhao et al. were within a close range given these expectations.

#### Modelling H&Y Transition Likelihood using Multi-state Hidden Markov Models in Continuous Time

Because real-world data is passively collected, data are not necessarily collected at fixed time points. As such, we modeled the likelihood of H&Y transitions after one year using multi-state Markov models. This analysis was performed using the “msm” package in R [6]. In this case, the continuously observed processes are patient interactions with the health system as part of their routine care. In HBS, the H&Y scores are available as structured data, directly recorded. In MGB, when the scores were recorded as part of normal patient care, we extracted the values associated with a particular patient visit date. This process allows for the modelling of transition likelihood in one year, which we use to represent an average or typical progression pattern.

#### Definitions of Health Care Utilization and Clinical Events

All definitions used to describe healthcare utilization and clinical events are derived from Beaulieu-Jones et al. where they are available in full detail in Supplemental Tables 1-3 of the prior work [4].

Within the MGB dataset, it is not always clear based on the discharge disposition if a patient is discharged to long-term care. We therefore analyzed discharge dispositions stratified by H&Y scores in two ways: 1) all discharge dispositions, 2) including only dispositions that are clearly self-care at home and long-term care in a skilled nursing facility (Table 1). Other discharge dispositions including “Home Health Service” were excluded because the level of care provided is not available.

## Data Availability

Authors do not have rights to share data, data from this study were made available from Mass General Brigham and the Harvard Biomarker Study. Authors will share all data upon approval from the data owner.

## Acknowledgements

We would like to thank the Parkinson’s community for participating in this study to make this research possible. We would like to acknowledge Cliona Molony and Sanofi Digital, for facilitating data access through the Sanofi Darwin platform to support this project. We would also like to thank all the participants and organizers of the Harvard Biomarker Study as well as the organizers of the Research Patient Data Registry at Mass General Brigham. We would especially like to thank Clemens Scherzer for his guidance and feedback throughout this work.

## Author Contributions

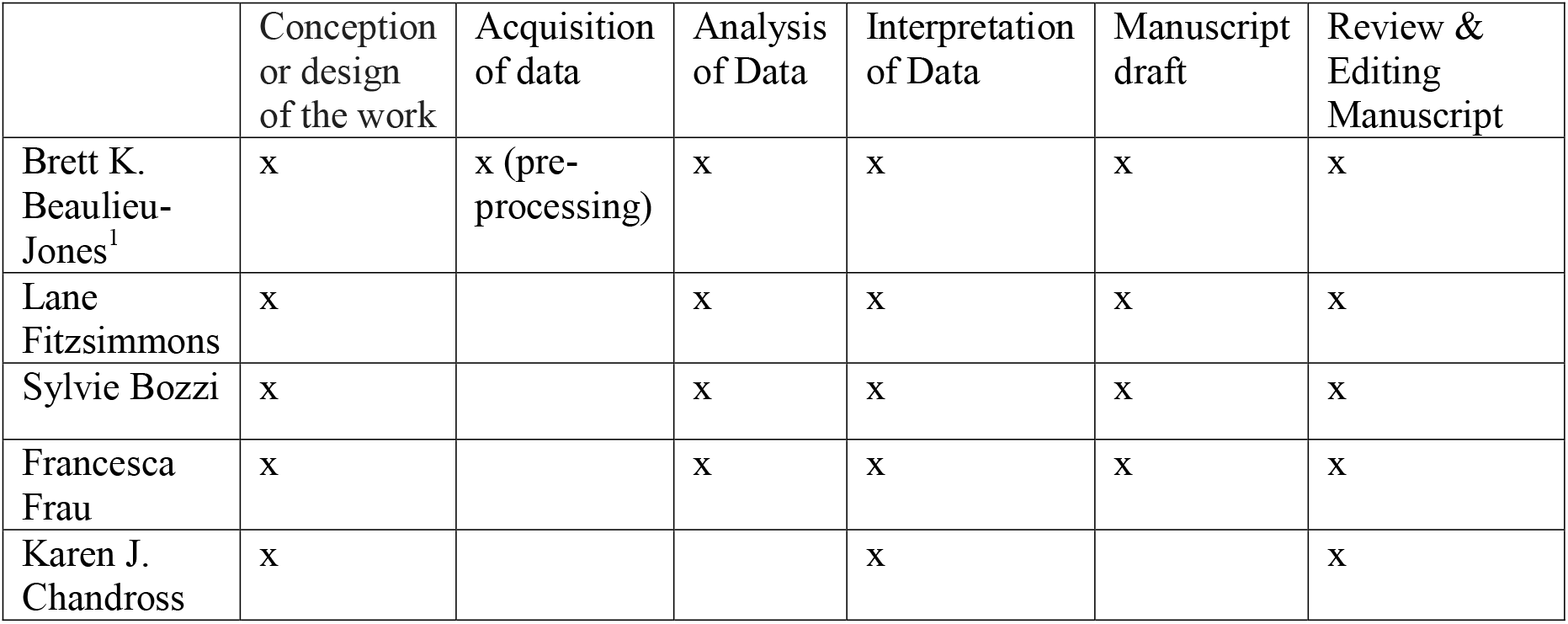

## Ethical Compliance Statement

The Mass General Brigham Institutional Review Board approved this study (protocol #2017P002452).

---

**Supplemental Table 1.** Modelled 12-month H&Y transition likelihood using Multi-state Hidden Markov Models of in Continuous Time.

| FROM<br>HY | TO HY |  |  |  |  |  |
| --- | --- | --- | --- | --- | --- | --- |
|  |  | 1 | 2 | 3 | 4 | 5 |
| 1 | HBS | 40.2% | 48.8% | 10.7% | 0.3% | 0.0% |
|  | MGB | 59.6% | 35.7% | 4.4% | 0.3% | 0.0% |
| 2 | HBS | 0.0% | 83.9% | 15.6% | 0.5% | 0.0% |
|  | MGB | 2.9% | 77.4% | 18.0% | 1.6% | 0.2% |
| 3 | HBS | 0.0% | 0.8% | 93.2% | 5.5% | 0.5% |
|  | MGB | 0.2% | 11.4% | 73.8% | 12.5% | 2.1% |
| 4 | HBS | 0.0% | 0.0% | 0.0% | 84.8% | 15.1% |
|  | MGB | 0.0% | 1.5% | 19.0% | 59.1% | 20.4% |
| 5 | HBS | 0.0% | 0.0% | 0.2% | 0.5% | 99.3% |
|  | MGB | 0.0% | 0.2% | 3.9% | 25.1% | 70.8% |

**Supplemental Table 2.** Cohort Characteristics for the four included data sources: HBS, MGB. Originally reported in Beaulieu-Jones et al. [4].

|  | HBS | MGB |
| --- | --- | --- |
| Total Population with PD | 935 | 22,949 |
| Population with coverage from PD onset (% Male) | 935 (67.02%) | 13,257 (57.81%) |
| Mean Age Onset (years) [25% and 75%] | 62.49<br>[56.00 – 70.00] | 72.86<br>[66.52 – 81.19] |
| Mean Years from Diagnosis to Enrollment [25%, 75%] | 3.66 [0.00 – 5.00] | NA |
| Mean Initial H&Y Stage if recorded within 1 year of diagnosis (MGB) or at enrollment (HBS) [25%, 50%, 75%] | 2.15 [2, 2, 2.5]<br>n=853 | 2.33 [2, 2, 3]<br>n=1,071 |
| Mean Length of Follow-up for de novo PD^ (years) [25%, 75% percentile] | 6.16 [2.00-9.00] | 3.84 [0.69 – 5.56] |

## References

1 Yang W, Hamilton JL, Kopil C, et al. Current and projected future economic burden of Parkinson’s disease in the U.S. NPJ Parkinsons Dis. 2020;6:15.

2 Hoehn MM, Yahr MD. Parkinsonism: onset, progression, and mortality. 1967. Neurology. 1998;50:318 and 16 pages following.

3 Zhao YJ, Wee HL, Au WL, et al. Corrigendum to “Selegiline use is associated with a slower progression in early Parkinson’s disease as evaluated by Hoehn and Yahr stage transition times” [Parkinsonism Relat Disord 17 (2011) 194–197]. Parkinsonism Relat Disord. 2011;17:299–300.

4 Beaulieu-Jones BK, Frau F, Bozzi S, et al. Disease progression strikingly differs in research and real-world Parkinson’s populations. npj Parkinson’s Disease. 2024;10:1–11.

5 Schrag A, Choudhury M, Kaski D, et al. Why do patients with Parkinson’s disease fall? A cross-sectional analysis of possible causes of falls. NPJ Parkinsons Dis. 2015;1:15011.

6 Jackson C. Multi-State Models for Panel Data: The msm Package for R. Journal of Statistical Software, Articles. 2011;38:1–28.

7 Jackson C. Multi-state modelling with R: the msm package. Cambridge, UK. 2007;1–53.

